# Current surgical practice for children born with a cleft lip and/or palate in the United Kingdom

**DOI:** 10.1101/2021.10.10.21264828

**Authors:** Matthew Fell, Alex Davies, Amy Davies, Shaheel Chummun, Alistair R.M. Cobb, Kanwalraj Moar, Yvonne Wren

## Abstract

**Background:** This study describes primary surgical reconstructions performed for children born with a cleft lip and/or palate in the United Kingdom.

**Methods:** Data were obtained from the Cleft Collective, a national longitudinal cohort study. Data forms completed at the time of surgery included details on timing, technique and adjuncts used during the operative period. Demographic data on participants were validated via parental questionnaires.

**Results:** Between 2015 and 2021, 1782 Cleft Collective surgical forms were included, relating to the primary reconstructions of 1514 individual children. The median age at primary cheiloplasty was 4.3 months. Unilateral cleft lips were reconstructed with an anatomical subunit approximation technique in 53%, whereas bilateral cleft lips were reconstructed with a broader range of eponymous techniques. Clefts of the soft palate were reconstructed at a median age of 10.3 months with an intravelar veloplasty in 94% cases. Clefts of the hard palate were reconstructed with a vomer flap in 84% cases in a bi-modal age distribution, relating to reconstruction carried out simultaneously with either lip or soft palate reconstruction. Antibiotics were used in 96% of cases, with an at-induction-only regimen used more commonly for cheiloplasties (p<0.001) and a 5–7day post-operative regime used more commonly for soft palatoplasties (p<0.001). Peri-operative steroids were used more commonly in palatoplasties than cheiloplasties (p<0.001) but tranexamic acid use was equivalent (p=0.73).

**Conclusion:** This study contributes to our understanding of current cleft surgical pathways in the United Kingdom and will provide a baseline for analysis of the effectiveness of utilised protocols.

## BACKGROUND

More than 1000 babies are born with cleft lip and/or palate (CLP) in the United Kingdom (UK) each year^1^ mirroring the global incidence of CLP approximated at 1/700 live births.^2^ The Clinical Standards Advisory Group (CSAG) in 1998 revolutionised cleft care in the UK by demonstrating superior aesthetic and functional outcomes were achieved in multidisciplinary cleft units treating high volumes of patients.^3^ The follow-up Cleft Care UK study in 2015 demonstrated that centralising care into 11 managed clinical networks and adhering to a national standard of care, including minimum numbers treated per surgeon, improved outcomes.^4–8^ The Oslo CLP surgical protocol; cheiloplasty and vomer flap repair of anterior hard palate at 3-6 months followed by soft palate repair at 6-9 months, greatly influenced UK cleft surgery pathways as it led to superior maxillary growth and dental arch relationships.^4–7^ Although the UK does not currently have a nationally agreed cleft surgery protocol, the national cleft quality indicators (published by the National Health Service) specify that cleft lips are repaired by 6 months and cleft palates by 13 months (in the absence of specific clinical reasons for delay).^8^

Determining current surgical practice for the primary reconstruction of CLP on a national level is a challenge. Previous efforts globally have tended to utilise surveys sent to cleft surgeons,^9,10^ which are limited by the reliance on the surgeon’s memory, risking that the recollected description may not accurately represent the surgeon’s approach to the range of heterogeneous cleft phenotypes. In the UK, the CSAG report had important ramifications for cleft research and led to the development of the Cleft Collective in 2012, a longitudinal cohort study set up to explore the causes, treatment and outcomes associated with CLP on individuals and their families.^11,12^ The aim of this study was to use Cleft Collective Birth Cohort Surgical Data to establish the current UK pathways for primary reconstructions of CLP in terms of operative timings, techniques and adjuncts used peri-operatively.

## METHODS

### Participants and Resource

Children born between 2014-2021 undergoing primary surgical reconstructions for CLP in the UK enrolled in the birth cohort of Cleft Collective Cohort studies were included.^12,13^ Secondary surgical interventions for speech, alveolar bone grafting, orthognathic or revisional procedures were excluded. The Cleft Collective resource comprises biological samples, speech audio recordings, medical and educational records and parent and child completed questionnaires. The resource is available for clinical and academic communities to use to address a range of cleft related research questions. More information on the study and how to access the dataset is available at http://www.bristol.ac.uk/cleft-collective/professionals/access/.

### Data collection and extraction

Details of surgical reconstruction were collected using the Cleft Collective Short Surgical Form (SSF) dataset, completed at the time of surgery by the operating surgeon, or delegated member of the team. The content and layout of the SSF was developed in discussion with UK cleft surgeons as a modification of the original Long Surgical Form, aiding completion by minimising the time required to complete it (see Supplementary Figure 1). Where the Long Surgical Form was completed, the truncated SSF dataset was extracted, but an analysis of the broader dataset was not included in this paper.

The SSF records patient demographics, cleft phenotype (LAHSHAL classification^14^ supplementary Table 1), date of surgery, reconstructive technique used (narrative) and use of perioperative antibiotics, tranexamic acid and steroids. Form data were uploaded into a database using scanning software in conjunction with manual checks. Cleft phenotype was validated with parent questionnaire and clinical data to verify accuracy.

### Data synthesis

Data were stratified by cleft phenotype as unilateral cleft lip only (UCL), bilateral cleft lip only (BCL) cleft palate only (CPO), unilateral cleft lip and palate (UCLP) and bilateral cleft lip and palate (BCLP). Narrative descriptions of surgical repair techniques were categorised independently by the first and second author using published techniques of reconstruction in cleft surgery.^15^ Differences were resolved through discussion to reach a consensus.

### Data analysis

The data were initially explored using descriptive and inferential statistics with medians (inter-quartile range (IQR)) used to describe continuous variables, and frequencies (percentages) used to describe categorical variables. The use of surgical adjuncts in primary cheiloplasty compared to soft palatoplasty was analysed using the Pearson chi-square test of independence. Odds ratios (OR), 95% confidence intervals (CIs) and P values were reported and interpreted as continuous measures of the strength of evidence against the null hypothesis.^16^ Due to the large sample size, we took a complete-case analysis approach and excluded missing data (See supplementary Table 2).^17^ Analysis was performed using the R Foundation for Statistical Computing Platform version 4.0.5 (http://www.R-project.org/).

### Ethical Approval

Ethical approval to establish the Cleft Collective Cohort Study was granted by the Southwest Central Bristol Ethics Committee (13/SW/0064). Global research and development (R&D) approval was provided by University Hospitals Bristol NHS Foundation Trust. Local R&D approvals were subsequently obtained from each National Health Service (NHS) Trust. National ethical approval (IRAS project ID 259689) to analyse this subset of data (Cleft Collective Project Number CC015) was approved by the NHS Health Research Authority.

## RESULTS

### Participants

From July 2015 to July 2021, the Cleft Collective received data for 1782 SSFs with completed cleft phenotype data relating to the primary cleft reconstructions of 1514 individual children. Demographic information is reported in Table 1. Of the 1782 forms, 849 had indicated a primary cheiloplasty (of which 312 had simultaneous repair of the hard palate) and 933 forms had indicated a primary palatoplasty (soft palate +-hard palate) had been performed. All 16 UK cleft surgical sites provided data contributing to this study.

**Table 1:** Demographic information of 1514 individual participants

| Individual variable | N | Percent |
| --- | --- | --- |
| **Gender** |  |  |
| Male | 875 | 58% |
| Female | 639 | 42% |
| **Cleft Phenotype** |  |  |
| Unilateral cleft lip only | 311 | 21% |
| Bilateral cleft lip only | 22 | 1% |
| Cleft palate only | 587 | 39% |
| Unilateral cleft lip and palate | 399 | 26% |
| Bilateral cleft lip and palate | 195 | 13% |
| **Recorded syndrome** |  |  |
| Yes | 123 | 8% |
| No | 1344 | 92% |

### Primary cleft lip reconstruction

Primary cheiloplasty was recorded on 849 forms with a median age of 4.3 months (IQR 3.6 to 5.4) (Figure 1) and did not differ markedly by cleft phenotype (see supplementary Table 3). Cheiloplasties were completed by the NHS threshold of 6 months in 700 (82%) cases.^8^

**Figure 1:**
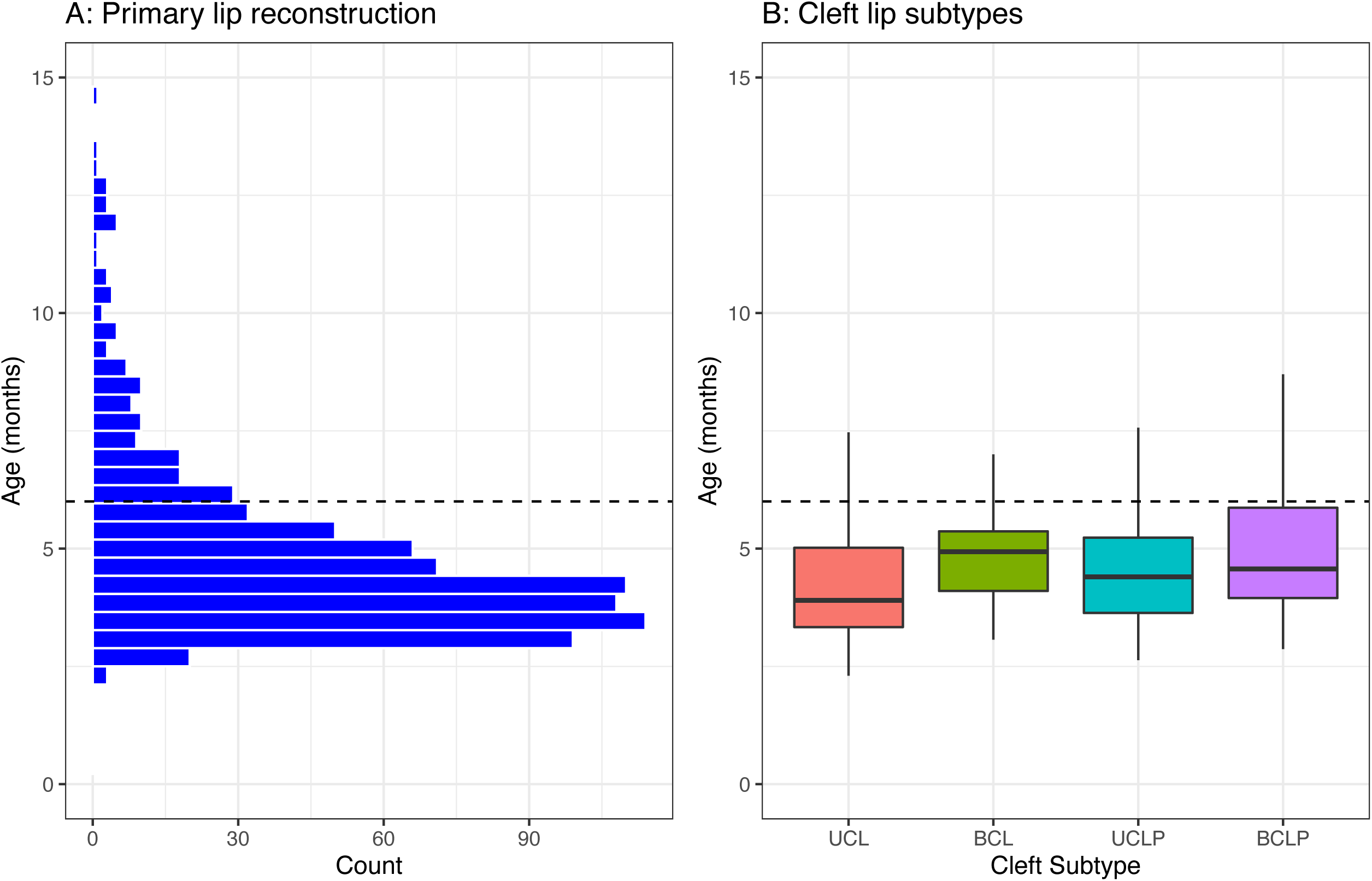
Age at primary lip reconstruction. A: Histogram shows distribution of age at all primary lip reconstruction and B: Box and whisker plot to show age of lip reconstruction by cleft subtype (unilateral cleft lip (UCL), bilateral cleft lip (BCL), unilateral cleft lip and palate (UCLP) and bilateral cleft lip and palate (BCLP)). The dashed line represents the UK National Health Service threshold age for lip reconstruction at 6 months of age.

Reconstructive techniques documented for UCL phenotype on 475 forms, showed the anatomical subunit approximation technique (often described as a Fisher^18^), reported in 252 (53%) cases (Figure 2), to be the most common, with a modified technique reported in an additional 28 (6%) cases. The second most common techniques were rotation advancement; described as a Millard^23^ in 71 (15%) cases, with modifications (including Mohler^19^, Noordhof^20^ and Cutting^21^) reported in a further 85 (18%). The least common technique was the inferior triangle reconstruction (described as a Tennison^22^ or Randall^23^) in 12 (3%) cases.

**Figure 2:**
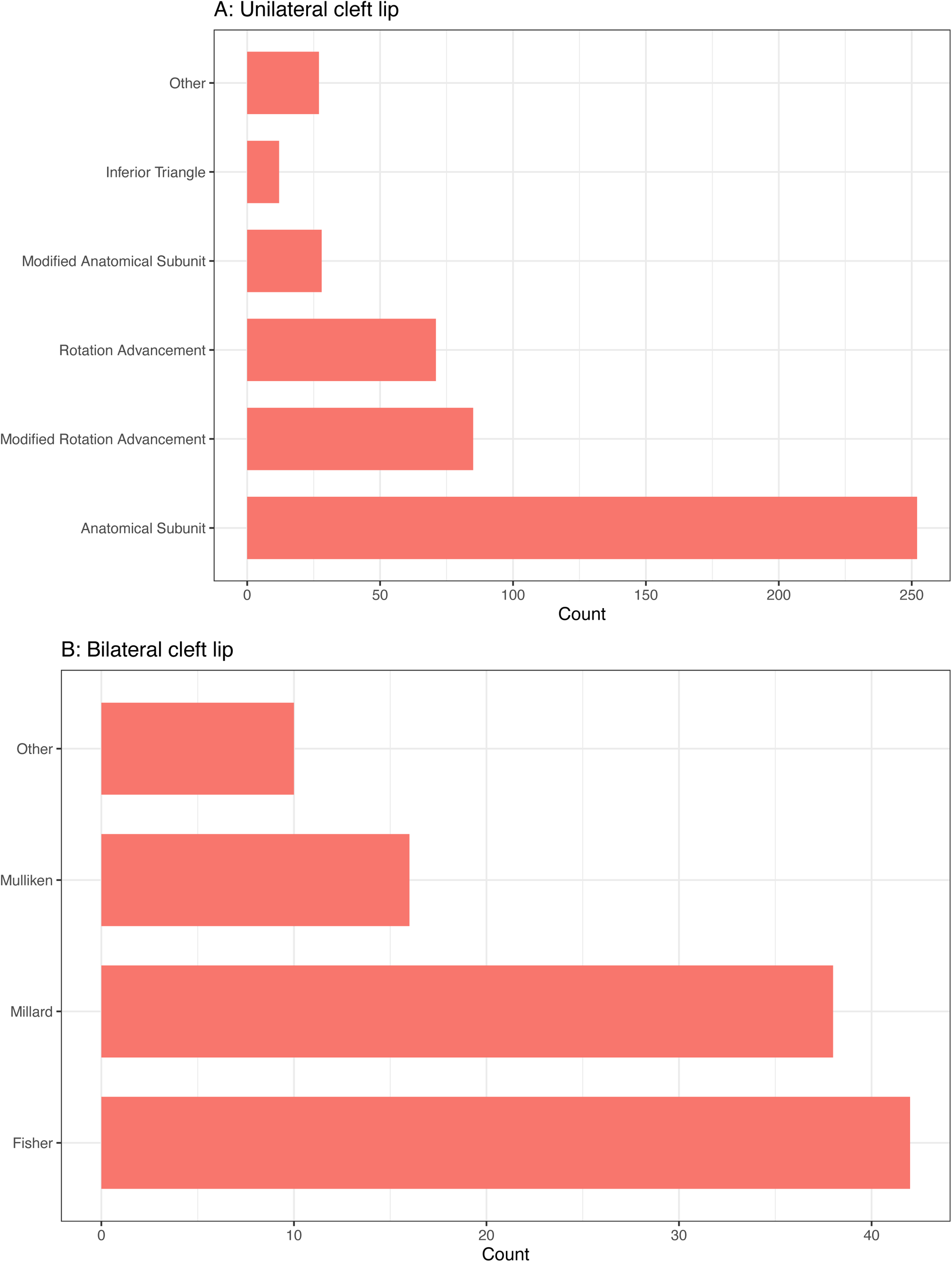
Named surgical reconstructive techniques for cleft lip. A: unilateral cleft lip reconstruction and B: bilateral lip reconstruction.

Reconstructive techniques for BCL phenotype on 142 forms documented a first stage lip adhesion in 36 cases (25%), 6 of which had additional forms submitted for subsequent definitive repair. For the 106 definitive BCL reconstructions, the most common techniques were eponymously named as Fisher^24^ in 42 cases (40%), Millard^25^ in 38 cases (36%) and Mulliken^26^ in 16 cases (15%).

### Primary palate reconstruction

Soft palate reconstructive techniques were recorded on 614 forms at a median age of 10.3 months (IQR 8.4 to 11.9) and occurred by the NHS threshold of 13 months in 526 (84%) cases (see Figure 3).^8^ The intravelar veloplasty (described as Sommerlad^27,28^) was the most used technique in 536 cases (94%) as shown in Figure 4. Relieving incisions were reported in combination with an intravelar veloplasty in 237 of the 536 cases (44%).

**Figure 3:**
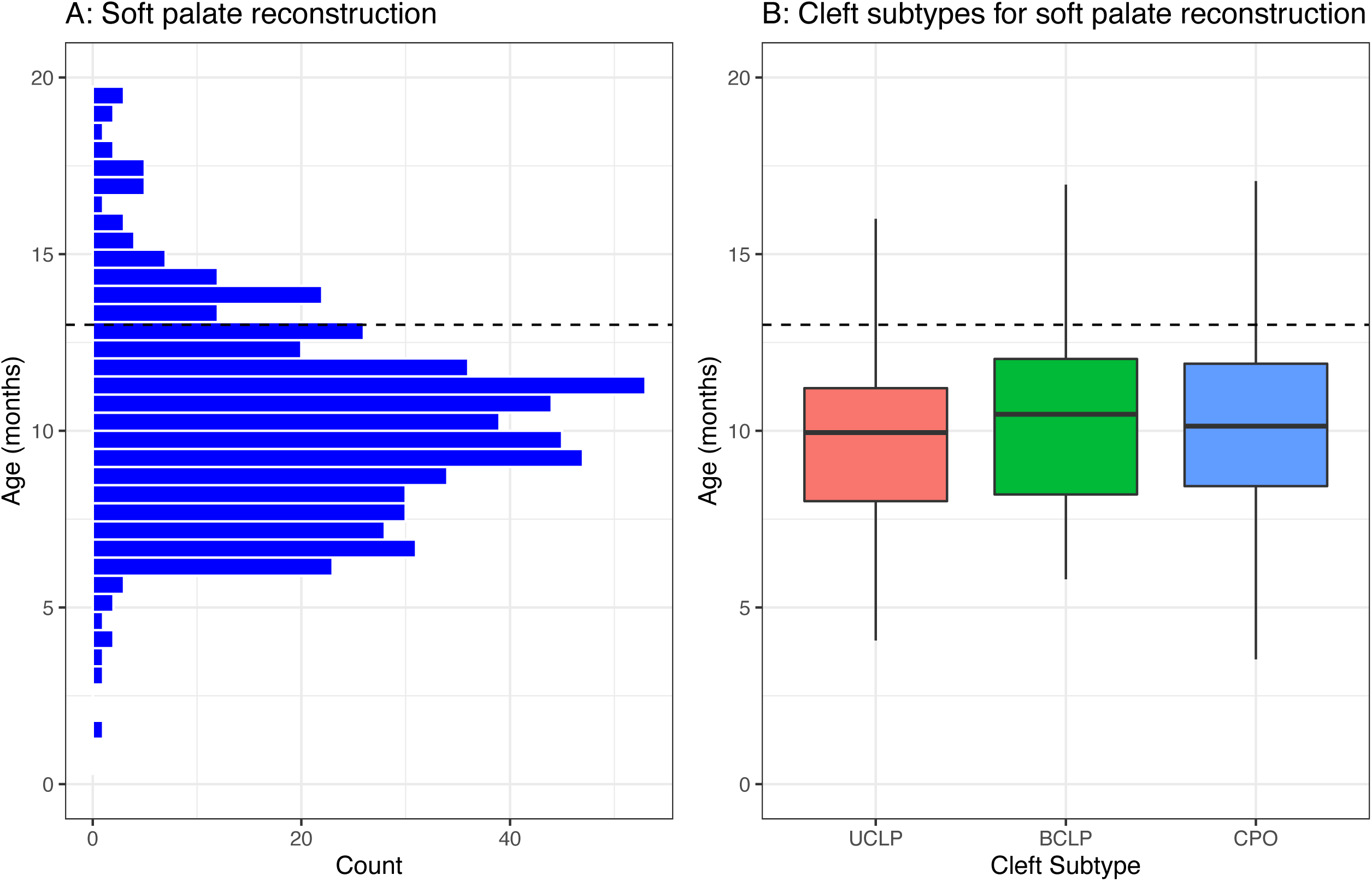
Age at primary soft palate reconstruction. A: Histogram shows distribution of age at all primary soft palate reconstructions and B: Box and whisker plot to show age of soft palate reconstruction by cleft subtype (unilateral cleft lip and palate (UCLP), bilateral cleft lip and palate (BCLP) and cleft palate only (CPO)). The dashed line represents the UK National Health Service threshold age for palatoplasty at 13 months of age.

**Figure 4:**
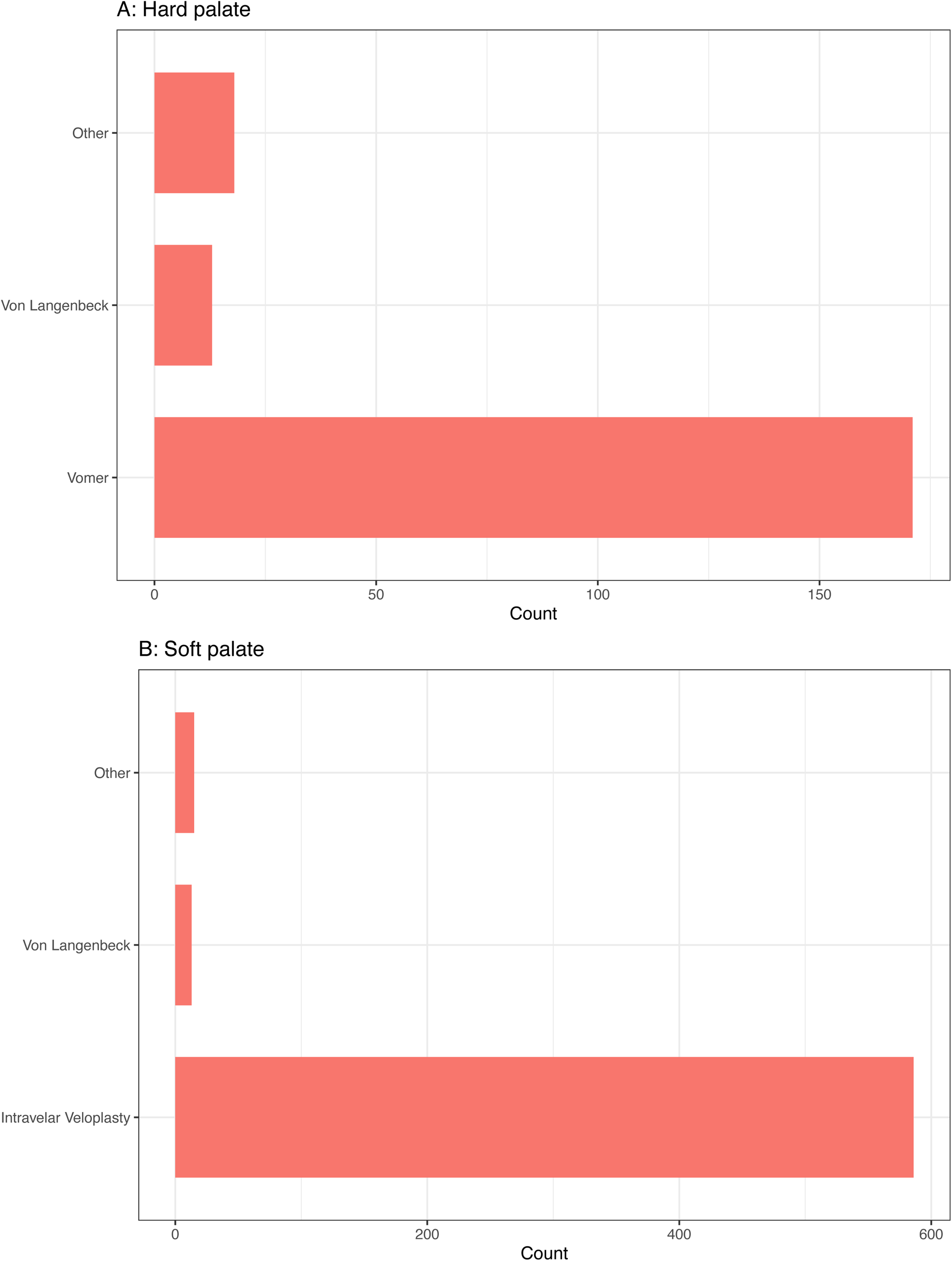
Named surgical reconstructive techniques for cleft palate A: hard palate reconstruction and B: soft palate reconstruction.

Hard palate reconstructive techniques were recorded on 203 forms; 136 (67%) were simultaneous with a primary cheiloplasty at median age of 4.4 months (IQR 3.7-5.4) as shown in Figure 5. Hard palate reconstruction exclusive of primary cheiloplasty occurred in 67 cases (33%), at a median age of 11.3 months (IQR 9.5-13.6). The vomer flap was the most used surgical technique in 171 cases (84%) as shown in Figure 4.

**Figure 5:**
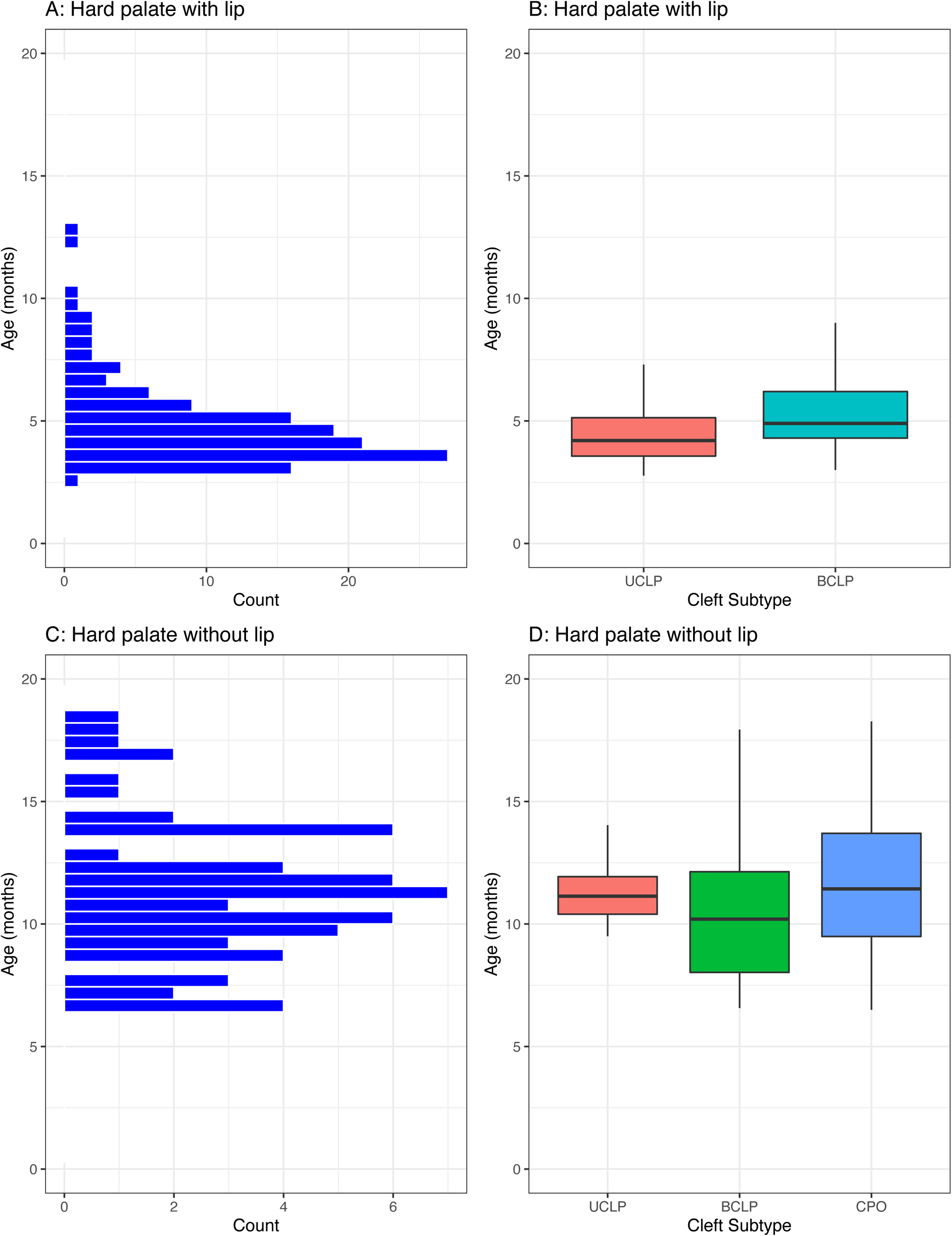
Age at primary hard palate reconstruction. A + B: distribution of age at reconstruction for hard palates when reconstructed simultaneously with a cleft lip, C+D: distribution of age when hard palate reconstructed without a lip repair. Cleft subtypes include unilateral cleft lip and palate (UCLP), bilateral cleft lip and palate (BCLP) and cleft palate only (CPO).

### Adjuncts

Of 1758 forms, antibiotics were used peri-operatively in 1694 (96%) primary cleft reconstructions with a variety of regimens used (See Table 2). There was strong evidence to suggest antibiotics were used more frequently at-induction-only for cheiloplasties compared to soft palatoplasties (OR 1.90, 95%CI 1.53-2.37; P<0.001) and a 5–7day post-operative course antibiotics was used more commonly in soft palatoplasty (OR 0.55, 95%CI 0.45-0.69; P<0.001). Co-amoxiclav was the most frequently used antibiotic, in 1311 cases (77%) (Supplementary Table 4).

**Table 2:** Antibiotic use within the sample overall with comparison made between cleft lip and soft palate reconstructions.

| Antibiotic regime | Overall<br>N(%) | Cleft Lip<br>N (%) | Soft Palate<br>N(%) | Odds Ratio<br>(95% CI) | X<sup>2</sup> | P value |
| --- | --- | --- | --- | --- | --- | --- |
| Number of forms<br>filled for antibiotics | 1758 | 841 | 610 | - | - | - |
| Antibiotics used at<br>all peri-operatively | 1694 (96%) | 806 (96%) | 593 (97%) | 0.52<br>(0.27, 1.03) | 3.63 | 0.057 |
| At induction only | 744 (42%) | 394 (47%) | 193 (32%) | 1.90<br>(1.53, 2.37) | 33.95 | <0.001 |
| Induction and post-op | 379 (22%) | 170 (20%) | 161(26%) | 0.71<br>(0.55, 0.90) | 7.67 | 0.006 |
| Post-op only | 571 (34%) | 24 (29%) | 239 (39%) | 0.63<br>(0.50, 0.78) | 17.27 | <0.001 |
| Use of any post-op<br>regimes: | 950 (54%) | 412 (49%) | 400 (66%) | 0.50<br>(0.40, 0.63) | 39.46 | <0.001 |
| Post-op (24 hours) | 367 (21%) | 181 (22%) | 140 (23%) | 0.81<br>(0.63, 1.03) | 3.00 | 0.083 |
| Post-op (5-7 days) | 583 (33%) | 245 (29%) | 260 (43%) | 0.55<br>(0.45, 0.69) | 28.36 | <0.001 |

Tranexamic acid was used in 810 of 1697 (48%) primary cleft reconstructions with weak evidence to show less common usage in cheiloplasty compared to soft palatoplasty (OR 0.96, 95%CI 0.78-1.19; P=0.73) (Table 3). Steroids were used in 1120 of 1663 (67%) primary cleft reconstructions with strong evidence to show less common usage in cheiloplasty compared to soft palatoplasty (OR 0.62, 95%CI 0.49-0.79; P<0.001).

**Table 3:** Adjuncts used at operative induction within the sample overall and comparison made between cleft lip and soft palate reconstructions.

| Adjunct at induction | Variable | Overall<br>N (%) | Cleft Lip<br>N (%) | Soft palate<br>N(%) | Odds Ratio<br>(95% CI) | X<sup>2</sup> | P<br>value |
| --- | --- | --- | --- | --- | --- | --- | --- |
| Tranexamic acid | Forms | 1697 | 806 | 588 |  |  |  |
|  | Use | 810 (47%) | 368 (46%) | 274 (47%) | 0.96<br>(0.78, 1.19) | 0.12 | 0.73 |
| Steroid | Forms | 1663 | 787 | 585 |  |  |  |
|  | Use | 1120 (67%) | 492 (63%) | 426 (73%) | 0.62<br>(0.49, 0.79) | 16.09 | <0.001 |

## DISCUSSION

This paper presents a unique insight into surgical pathways used in the UK for the primary reconstruction of CLP, using contemporaneous data collection at the point of care on a case-by-case basis. Previous national efforts have relied on retrospective surveys sent to surgeons to describe their practice.^10,29–37^

### Timing

The timing of cleft lip reconstruction (IQR 4 to 5 months) and soft palate reconstruction (IQR 8 to 12 months) is similar to previous UK reports,^9,35,38^ and does not appear to have altered with centralisation of care over the last three decades. Where primary reconstruction was delayed beyond the NHS threshold ages, the SSF data could not elucidate the reason, but has been reported elsewhere to occur more commonly in children with co-morbidities.^39^ We would hope the baseline data from this study in conjunction with outcome data that is being collected will contribute to the debate around optimal timing for palatal surgery in relation to critical speech development,^43^ psychological and growth outcomes.^44-46^ The bimodal timing of hard palate closure at 4-5 months and 10-14 months confirms hard palates commonly being reconstructed in combination with a lip or soft palate and rarely as a stand-alone procedure.

### Cleft Lip Techniques

The anatomical subunit approximation as described in 2005 by Fisher^18^ was the most used technique for UCL in 53% cases. This represents a change in the UK from 1988, where the favoured approach was the rotation-advancement.^35^ The anatomical subunit technique aims to leave a scar on lip subunit boundaries, achieving lip lengthening via the Rose-Thompson effect, and in most cases a small triangle above the white roll.^15^ The rotation-advancement technique, originally described by Millard^40^ in the 1950s, involves a curvilinear incision on the medial lip element to provide rotation and a triangular flap on the lateral element to provide advancement.^15^ Modifications of the rotation-advancement were used more frequently in this study compared to the original Millard technique itself. Rotation-advancement modifications recorded on the surgical form included Mohler,^19^ which extends the incision into the columella to enhance the lip lengthening and Noordhof^20,41^ which adds a small triangle above the white roll and a laterally based vermillion triangle.

Globally the rotation-advancement technique, and its modifications, is reported to be the most used for UCL repair.^29,31–33^ It is described as a “cut as you go” technique, whereas the anatomical subunit has been termed a “measure twice, cut once” technique.^42^ Given its relatively recent introduction in 2005, the popularity of the anatomical subunit reconstruction in the UK is quite remarkable. The reason for its adoption cannot be ascertained from data in our study but a surgeon’s choice of UCL repair technique has been previously described to be a hybrid of training, experience and imagination.^43^

A first stage lip adhesion was recorded in 25% cases of BCL in this study, which is higher than the 11% of surgeons reported to use a staged approach in the USA, where pre-surgical orthopaedics are commonly utilised.^30^ There was greater variation in techniques used for definitive BCL reconstruction compared to UCL in this study. The three eponymous techniques of Fisher^24^, Millard^25^ and Mulliken^26^ all share a common principle of recruiting tissue from the lateral lip elements to recreate the Cupid’s bow and vermillion across the prolabium.^44^ The Manchester^45^ repair uses vermillion native to the prolabium but was not reported in our cohort, in contrast to its use by 12% of US surgeons.^30^ The use of Millard and Mulliken techniques for BCL reconstruction does not come as a surprise due to the longevity and volume of the technique descriptions in the literature.^44^ The common use of Fisher’s approach to the BCL suggests a rapid adoption following its more recent description in 2009.^24^

### Cleft Palate Techniques

Clefts of the soft palate were reconstructed almost exclusively using the intravelar veloplasty (94% cases) and this is a change from 1988 in the UK, where the most used technique was the straight-line closure described by Von-Langenbeck.^35^ The intravelar veloplasty, described initially in 1970 by Kriens^46^ was popularised in the UK by Sommelad following his publications in 2003^27,28^ on the radical method of muscle retro-positioning and use of the operating microscope. The von Langenbeck^47^ technique of incisions along the medial cleft margins with lateral incisions can be used in combination with an intravelar veloplasty.^48^ The UK appears relatively unique in its sole adoption of the intravelar veloplasty for soft palate reconstruction, as, in comparison, the double-opposing z-plasty described by Furlow^49^ often has equivalent popularity elsewhere.^30,31,34^

Clefts of the hard palate were reconstructed with a vomer flap in 84% cases, as first described by Pichler.^50^ A vomer flap can be used variably; performed simultaneously with a cheiloplasty in a single-layered closure of the nasal mucosa, or in conjunction with a soft palate repair where a vomer flap(s) will likely reconstruct the nasal mucosa within a two-layered closure. Asher-McDade and Shaw (1990) noted an increase in its popularity^35^ for anterior hard palate reconstruction in the UK. Globally, surgeons have reported using a variety of techniques for reconstruction of the hard palate which commonly include the pushback technique described by Veau-Wardill-Kilner^51–53^ and the Bardach^54^ two-flap technique.^10,31,33,34^ The sequence of reconstructing the lip and anterior hard palate first followed by the soft palate later is in accordance with the Oslo protocol and has been previously described as the most commonly used in the UK.^7,9^ There is not yet global consensus on the optimal protocol for a cleft of the lip and palate.^15,55–57^

### Adjuncts

Reconstructions of CLP are classified as clean-contaminated surgical procedures with gram-positive organisms reported at the surgical site in multiple studies.^58,59^ Although little evidence or guidance is available to support the use of prophylactic antibiotics in elective cleft surgery, prophylactic intravenous antibiotics were used in 97% of cases within this cohort, no change from a survey of UK cleft surgeons in 2004, which reported most prescribing prophylactic antibiotics.^58^ We found strong evidence to suggest UK cleft surgeons more commonly prescribe a 5-7day post-operative course of antibiotics following soft palatoplasty than cheiloplasty, possibly reflecting concerns of intra-oral infection leading to palatal fistulae. A randomised trial conducted in India compared two groups of cleft patients, being given a single pre-operative dose of antibiotics or an additional five days of post-operative antibiotics. The study concluded, in spite of high loss to follow up, there was no difference in the early complication rate. Some evidence suggested a reduced fistula rate if post-operative antibiotics were given in this setting.^60^

Tranexamic acid was used intravenously on induction in half of the cases in this cohort, with weak evidence to show any difference in use between soft palatoplasty and cheiloplasty. The efficacy of tranexamic acid in palatoplasty has been evaluated in two randomised trials with one concluding that tranexamic acid markedly improved surgeon reported satisfaction^61^ and the other reporting no difference in in the amount of blood loss.^62^

Steroids were used intravenously on induction in 67% cases in this cohort, with strong evidence to show use more commonly for soft palatoplasty than cheiloplasty and this may be due to the perceived benefit to the child in terms of reducing post-operative airway swelling and improving oral intake.^63^ A randomised study to compare 0.25mg/kg steroid on induction to placebo in palatoplasty reported reductions in airway distress and post-operative fever associated with the steroid group.^64^ A more recent correspondence described a randomised study of 0.5mg/kg IV dexamethasone on induction in palatoplasty and reported an improvement in oral intake in the steroid group.^65^

### Strengths and Limitations

This study describes the operative pathways used within the centralised UK cleft services without the risk of recall bias associated with surgeon questionnaires previously used in this field. The large number of participants, inclusion of all CLP subtypes and contributions from all cleft centres in the UK ensures this cohort is broadly representative of UK cleft practices.

Reporting bias should be considered a limitation, as one description of an operation by name might not match either the original description or another’s interpretation of the same operation. There are limitations regarding the extent to which the intra-operative decision-making process and the techniques used by the cleft surgeon can be gleaned from SSF data. The principles involved with the reconstruction of an orofacial cleft include planning, wide surgical release and reconstruction of component parts in layers.^42^ The name of an operation gives an indication of the planning stage but no information on the other principle elements of the reconstruction. The SSF did not facilitate documentation of the approach to the nose in primary cheiloplasties and this is an important part of the reconstruction.^66^

## CONCLUSION

This paper presents a unique insight into UK cleft surgical pathways, providing information on the timing of surgical interventions, the operative techniques, and the use of surgical adjuncts. The provision of cleft care in the UK was changed significantly with the implementation of centralisation 20 years ago. Although the timing of primary CLP surgery has not changed, there has been a paradigm shift in the surgical techniques used. The anatomical subunit approximation technique has gained in popularity for UCL repair and the intravelar veloplasty is by far the most commonly used method for soft palate repair. The findings from this report can be used in conjunction with growing outcome data from national registries to analyse the effectiveness of the cleft protocols in use within the UK.

## Data Availability

All data produced in the present work are contained in the manuscript

## ACKNOWLEDGEMENTS

This publication involves data derived from independent research funded by The Scar Free Foundation (REC approval 13/SW/0064). We are grateful to the families who participated in the study, the UK NHS cleft teams, and The Cleft Collective team, who helped facilitate the study. The views expressed in this publication are those of the author(s) and not necessarily those of The Scar Free Foundation or The Cleft Collective Cohort Studies team.

## Supplementary data

**Supplementary Figure 1:**
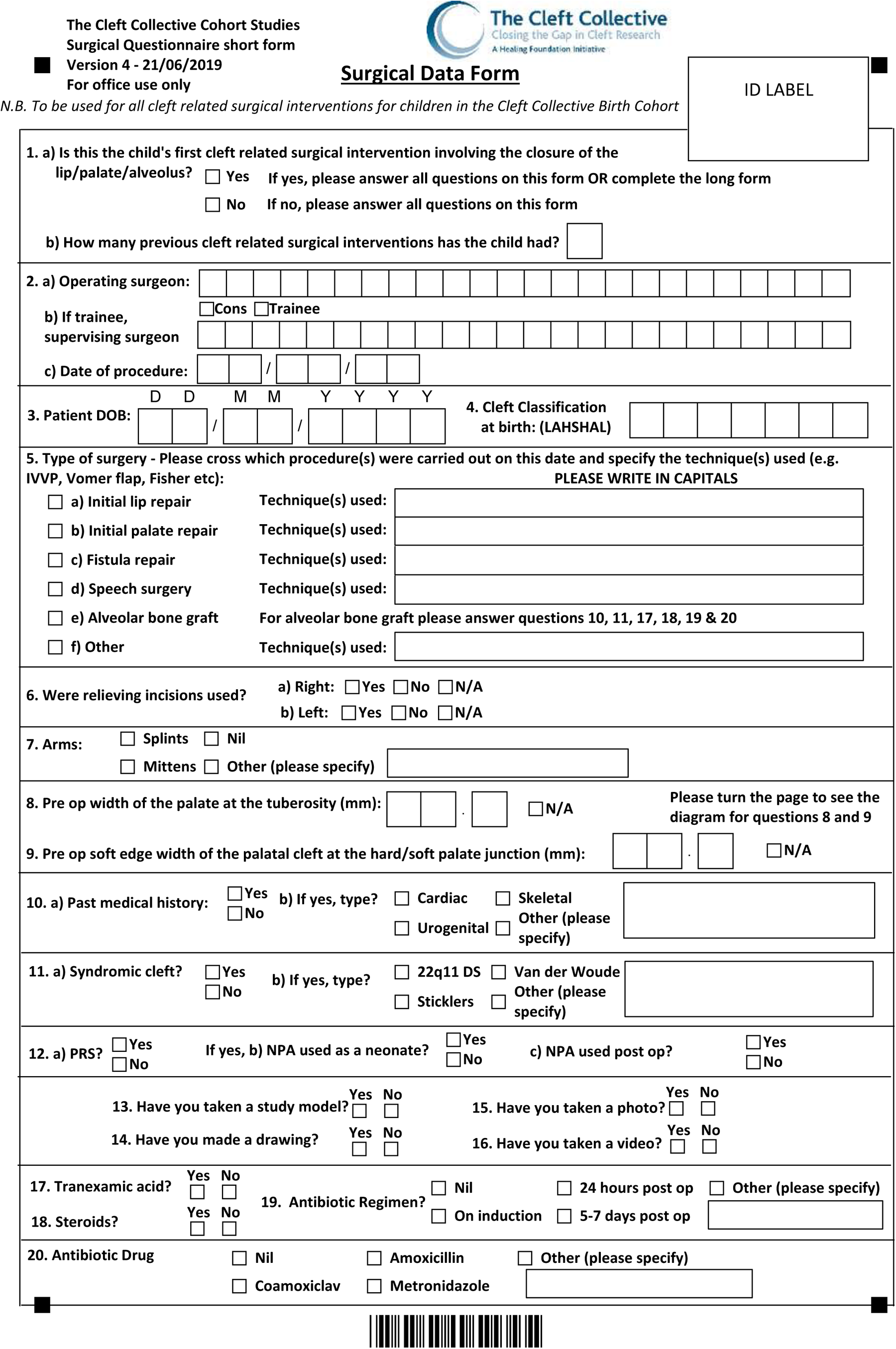
Cleft Collective Short Surgical Form

**Supplementary Table 1:** Stratification by cleft phenotype was performed according to the LAHSHAL classification. Surgeons noted the LAHSHAL on the Cleft Collective Surgical from and this was verified with clinical data to check for accuracy. We were not able to differentiate between incomplete and complete clefts in this study, therefore code for incompletes (denoted by non-capitalised letters) was included.

| Cleft phenotype | LAHSHAL Classification |
| --- | --- |
| **UCL** | L..... |
|  | .....L |
|  | LA..... |
|  | .....AL |
| **BCL** | L.....L |
|  | LA....L |
|  | L....AL |
|  | LA...AL |
| **CPO** | ...S... |
|  | ..HS... |
|  | ...SH.. |
|  | ..HSH.. |
| **UCLP** | LAHS... |
|  | ...SHAL |
|  | LA.S... |
|  | ...S.AL |
|  | L..S... |
|  | ...S..L |
| **BCLP** | LAHSHAL |

**Supplementary Table 2:** Missing data from the 1782 Cleft Collective Short Surgical Form dataset included in this study

| Variable | Number of missing data | Percent of missing data |
| --- | --- | --- |
| Cleft phenotype | 0 | 0% |
| Sex | 0 | 0% |
| Syndrome | 57 | 3% |
| Age at operation | 64 | 4% |
| Antibiotic | 24 | 1% |
| Tranexamic Acid | 85 | 5% |
| Steroids | 119 | 7% |

**Supplementary Table 3:** Age in months at primary cleft reconstruction stratified by cleft phenotype. Cleft phenotypes are stratified according to the LAHSHAL classification into All (any cleft lip and/or palate), UCL (unilateral cleft lip only), BCL (bilateral cleft lip only), CPO (cleft palate only), UCLP (unilateral cleft lip and palate) and BCLP (bilateral cleft lip and palate). Age in months is reported in median, lowest age, highest age, first quartile (1^st^Q), third quartile (3^rd^Q) and interquartile range (IQR)

| Operation | Cleft phenotype | Median age | Lowest age | Highest age | 1<sup>st</sup>Q | 3<sup>rd</sup>Q | IQR |
| --- | --- | --- | --- | --- | --- | --- | --- |
| Primary cheiloplasty | All | 4.3 | 2.3 | 72.8 | 3.6 | 5.4 | 1.8 |
|  | UCL | 4.0 | 2.3 | 72.0 | 3.4 | 5.2 | 1.8 |
|  | BCL | 4.9 | 3.1 | 72.8 | 4.1 | 5.5 | 1.4 |
|  | UCLP | 4.4 | 2.6 | 42.9 | 3.6 | 5.2 | 1.6 |
|  | BCLP | 4.6 | 2.9 | 21.7 | 4.0 | 6.0 | 2 |
| Primary hard palatoplasty *with* cheiloplasty | All | 4.4 | 2.8 | 12.7 | 3.7 | 5.4 | 1.7 |
|  | UCLP | 4.2 | 2.8 | 12.1 | 3.6 | 5.1 | 1.5 |
|  | BCLP | 4.9 | 3.0 | 12.7 | 4.3 | 6.2 | 1.9 |
| Primary hard palatoplasty *without* cheiloplasty | All | 11.2 | 6.5 | 24.4 | 9.5 | 13.6 | 4.1 |
|  | CPO | 11.5 | 6.5 | 24.4 | 9.5 | 13.9 | 4.4 |
|  | UCLP | 11.1 | 7.3 | 14.1 | 10.4 | 11.9 | 1.5 |
|  | BCLP | 10.3 | 6.6 | 22.9 | 8.3 | 13.3 | 5 |
| Primary soft palatoplasty | All | 10.3 | 1.8 | 52.7 | 8.4 | 11.9 | 3.5 |
|  | CPO | 10.3 | 1.8 | 52.7 | 8.5 | 12.0 | 3.5 |
|  | UCLP | 10.0 | 3.0 | 33.2 | 8.2 | 11.3 | 3.1 |
|  | BCLP | 11.3 | 5.1 | 27.2 | 8.3 | 12.5 | 4.2 |

**Supplementary Table 4:** Antibiotics prescribed in the peri-operative period for primary reconstruction of a cleft lip and/or palate.

| Antibiotic | N | Percent |
| --- | --- | --- |
| Prescribed | 1707 | 100% |
| Co-amoxicillin | 1311 | 77% |
| Benzylpenicillin | 143 | 8% |
| Amoxicillin | 20 | 1% |
| Metronidazole | 10 | 0.5% |
| Other | 223 | 13% |

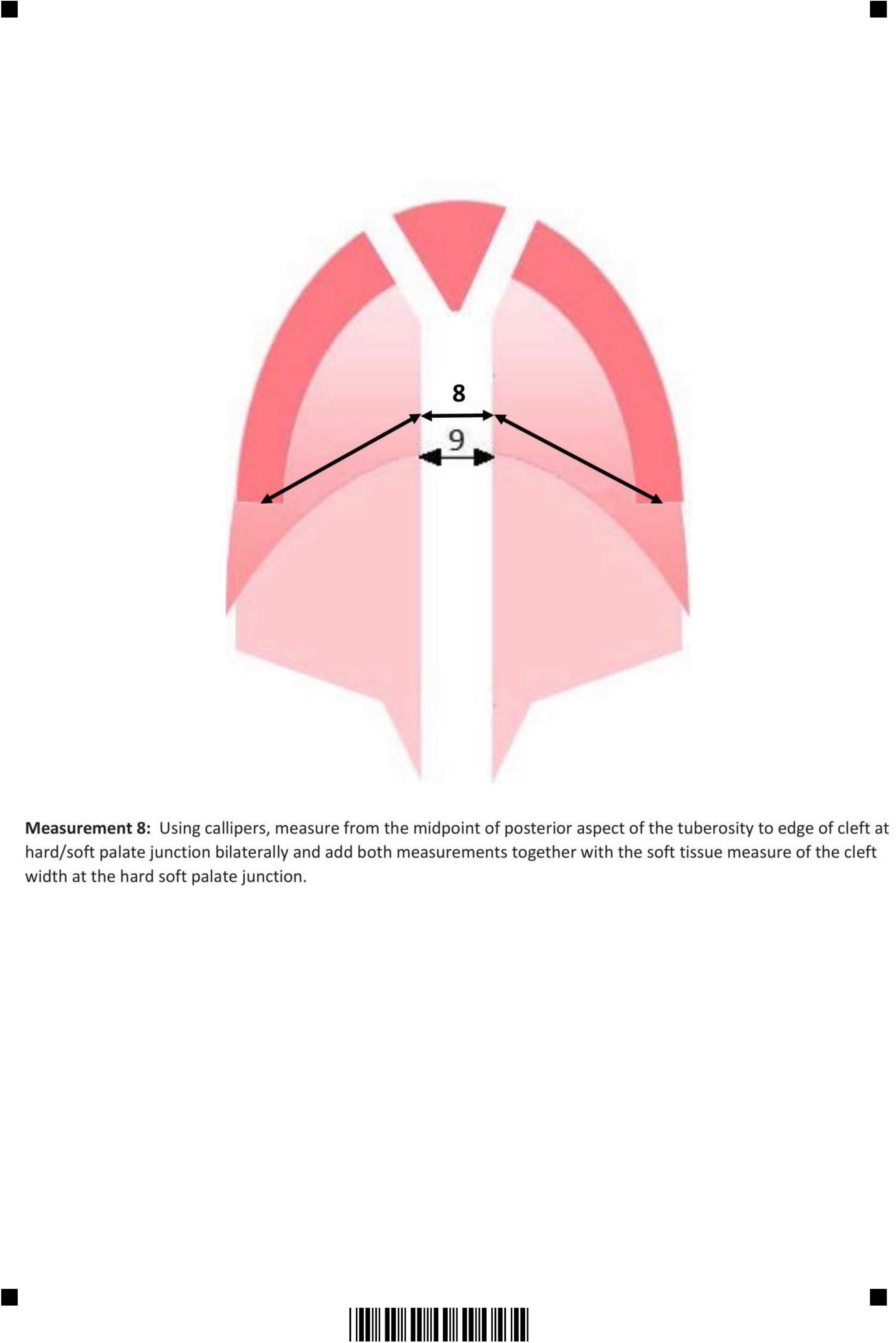

## REFERENCES

1. The Cleft Registry and Audit Network (CRANE). Crane Database 2020 Annual Report. Available at: https://www.crane-database.org.uk/reports/crane-database-2020-annual-report/. Accessed September 30, 2021.

2. Mossey PA, Little J, Munger RG, Dixon MJ, Shaw WC. Cleft lip and palate. Lancet. 2009;374:1773–1785.

3. Bearn D, Mildinhall S, Murphy T, Murray JJ, Sell D, Shaw WC et al. Cleft lip and palate care in the United Kingdom - The Clinical Standards Advisory Group (CSAG) Study. Part 4: Outcome comparisons, training, and conclusions. Cleft Palate Craniofac J. 2001;38:38–43.

4. Mars M, Asher-McDade C, Brattstrom V, Dahl E, McWilliam J, Molsted K et al. A six-center international study of treatment outcome in patients with clefts of the lip and palate: Part 3. Dental arch relationships. Cleft Palate Craniofac J. 1992;29:405–408.

5. Mølsted K, Asher-Mcdade C, Brattström V, Dahl E, Mars M, McWilliam J et al. A Six-Center International Study of Treatment Outcome in Patients with Clefts of the Lip and Palate: Part 2. Craniofacial Form and Soft Tissue Profile. Cleft Palate Craniofac J. 1992;29:398–404.

6. Pigott RW, Albery EH, Hathorn IS, Atack NE, Williams A, Harland K et al. A comparison of three methods of repairing the hard palate. Cleft Palate Craniofac J. 2002;39:383–391.

7. Colbert SD, Green B, Brennan PA, Mercer N. Contemporary management of cleft lip and palate in the United Kingdom. Have we reached the turning point? Br J Oral Maxillofac Surg. 2015;53:594–598.

8. National Health Service (NHS) England. Cleft Lip and/or Palate Quality Dashboard 2018/2019. Available at: https://www.england.nhs.uk/wp-content/uploads/2018/03/clp-metric-definitions-2018-19.pdf. Accessed September 30, 2021.

9. Slator R, Perisanidou LI, Waylen A, Sandy J, Ness A, Wills AK. Range and timing of surgery, and surgical sequences used, in primary repair of complete unilateral cleft lip and palate: The Cleft Care UK study. Orthod Craniofac Res. 2020;23:166–173.

10. Katzel EB, Basile P, Koltz PF, Marcus JR, Girotto JA. Current surgical practices in cleft care: Cleft palate repair techniques and postoperative care. Plast Reconstr Surg. 2009;124:899–900.

11. Seifert M, Davies A, Harding S, McLeod S, Wren Y. Intelligibility in 3-Year-Olds With Cleft Lip and/or Palate Using the Intelligibility in Context Scale: Findings from the Cleft Collective Cohort Study. Cleft Palate Craniofac J. 2021;58:1178–1189.

12. The Cleft Collective. Available at: www.cleftcollective.org.uk. Accessed September 30, 2021

13. Stock NM, Humphries K, Pourcain BS, Bailey M, Persson M, Ho KM et al. Opportunities and challenges in establishing a cohort study: An example from cleft lip/palate research in the United Kingdom. Cleft Palate Craniofac J. 2016;53:317–325.

14. Kriens O. Lahshal: A concise documentation system for cleft lip, alveolus and palate diagnoses. In: Kriens O, ed. What Is Cleft Lip and Palate? A Multidisciplinary Update Workshop. New York: Thieme Medical Publishers; 1987:30–34

15. Fisher DM, Sommerlad BC. Cleft lip, cleft palate, and velopharyngeal insufficiency. Plast Reconstr Surg. 2011;128:342–360.

16. Greenland S, Senn SJ, Rothman KJ, Carlin JB, Poole C, Goodman SN et al. Statistical tests, P values, confidence intervals, and power: a guide to misinterpretations. Eur J Epidemiol. 2016;31:337–350

17. Karahalios A, Baglietto L, Carlin JB, English DR, Simpson JA. A review of the reporting and handling of missing data in cohort studies with repeated assessment of exposure measures. BMC Med Res Methodol. 2012;12:96.

18. Fisher DM. Unilateral cleft lip repair: An anatomical subunit approximation technique. Plast Reconstr Surg. 2005;116(1):61–71.

19. Mohler LR. Unilateral cleft lip repair. Plast Reconstr Surg. 1987;80:511–517

20. Noordhoff MS. Reconstruction of vermilion in unilateral and bilateral cleft lips. Plast Reconstr Surg. 1984;73:52–60.

21. Cutting CB, Dayan JH. Lip Height and Lip Width after Extended Mohler Unilateral Cleft Lip Repair. Plast Reconstr Surg. 2003;111:17–23.

22. Tennison CW. The repair of the unilateral cleft lip by the stencil method. Plast Reconstr Surg. 1952;9:115–120

23. Randall P. A triangular flap operation for the primary repair of unilateral clefts of the lip. Plast Reconstr Surg. 1959;23:331–347.

24. Fisher D. Bilateral Cleft Lip. In: Guyuron B, Eriksson E, Persing JA, Chung KC, Disa J, Gosain A, Kinney B, Rubin JP eds. Plastic Surgery: Indications and Practice. Saunders; 2009:493–504.

25. Millard DR, Cassisi A, Wheeler JJ. Designs for correction and camouflage of bilateral clefts of the lip and palate. Plast Reconstr Surg. 2000;105:1609–1623.

26. Mulliken JB. Repair of bilateral complete cleft lip and nasal deformity -State of the art. Cleft Palate Craniofac J. 2000;37:342–347.

27. Sommerlad BC. A technique for cleft palate repair. Plast Reconst Surg. 2003;112:1542–1548.

28. Sommerlad BC. The use of the operating microscope for cleft palate repair and pharyngoplasty. Plast Reconstr Surg. 2003;112:1540–1541.

29. Sitzman TJ, Girotto JA, Marcus JR. Current surgical practices in cleft care: Unilateral cleft lip repair. Plast Reconstr Surg. 2008;121:261–270.

30. Tan SPK, Greene AK, Mulliken JB. Current surgical management of bilateral cleft lip in North America. Plast Reconstr Surg. 2012;129:1347–1355.

31. Abulezz T, Elsherbiny A, Mazeed A. Management of cleft lip and palate in Egypt: A National survey. Indian J Plast Surg. 2018;51:290–295.

32. Lee TJ, Kim ST. A survey of cleft lip and palate management taught in training programs in Korea. Cleft Palate Craniofac J. 2003;40:80–83.

33. Gopalakrishna A, Agrawal K. A status report on management of cleft lip and palate in India. Indian J Plast Surg. 2010;43:66–75.

34. Franco D, Gonçalves LF, Franco T. Management of cleft lip and palate in Brazil. Scand J Plast Reconstr Surg Hand Surg. 2003;37:272–276.

35. Asher-McDade C, Shaw WC. Current cleft lip and palate management in the United Kingdom. Br J Plast Surg. 1990;43:318–321.

36. Pigott RW. Organisation of cleft lip and palate services - results of a questionnaire. Br J Plast Surg. 1992;45:385–387.

37. Brennan PA, Macey-Dare LV, Flood TR, Markus AF, Uppal R. Cleft lip and palate management by U.K. Consultant oral and maxillofacial surgeons: A national survey. Cleft Palate Craniofac J. 2001;38:44–48.

38. Fitzsimons KJ, Mukarram S, Copley LP, Deacon SA, van der Meulen JH. Centralisation of services for children with cleft lip or palate in England: A study of hospital episode statistics. BMC Health Serv Res. 2012;12:148.

39. Butterworth S, Rivers C, Fullarton M, Murphy C, Beale V, Neil-Dwyer J et al. A Closer Look at Delayed Primary Cleft Surgery and Unrepaired Cleft Lip and/or Palate in 5 UK Cleft Centers. Cleft Palate Craniofac J. 2021. Epub ahead of print.

40. Millard DR Jr. Complete unilateral clefts of the lip. Plast Reconstr Surg Transplant Bull. 1960;25:595–605.

41. Samuel Noordhoff M, Chen YR, Chen KT, Hong KF, Lo LJ. The surgical technique for the complete unilateral cleft lip-nasal deformity. Oper Tech Plast Reconstr Surg. 1995;2:167–174.

42. Tse R. Unilateral cleft lip: Principles and practice of surgical management. Semin Plast Surg. 2012;26:145–155.

43. Thomson H. Unilateral cleft lip repair. Oper Tech Plast Reconstr Surg. 1995:2:175-181

44. Zhang JX, Arneja JS. Evidence-Based Medicine: The Bilateral Cleft Lip Repair. Plast Reconstr Surg. 2017;140(1). 152e-165e.

45. Manchester WM. The repair of bilateral cleft lip and palate. Br J Surg. 1965;52:878–882.

46. Kriens OB. Fundamental anatomic findings for an intravelar veloplasty. Cleft Palate J. 1970;7:27–36

47. Von Langenbeck B. Operation der angeborenen totalen Spaltung des harten Gaumens nach einer neuen Methode. Dtsch Klin. 1861;8:231.

48. Hopper RA, Tse R, Smartt J, Swanson J, Kinter S. Cleft palate repair and velopharyngeal dysfunction. Plast Reconstr Surg. 2014;133:852–864.

49. Furlow LT. Cleft palate repair by double opposing z-plasty. Plast Reconstr Surg. 1986;78:724–738.

50. Pichler H. Operationen der angeborenen Lippen-Kiefer-Gaumenspalten. Wien Klin Wochenschr. 1934;47:70–72.

51. Veau V, Ruppie C. Anatomie chirurgicale de la division palatine: Considerations operatoires. Rev Chir. 1922;20:1–30.

52. Wardill W. Technique of operation for cleft lip and palate. Br J Surg. 1937;25:117–130.

53. Kilner T. Cleft lip and palate repair technique. In: Maingot R, ed. Postgraduate Surgery. Vol 3. Medical Publishers; 1937:3800–3827.

54. Bardach J. Two-Flap palatoplasty: Bardach’s technique. Oper Tech Plast Reconstr Surg. 1995;2:211–214.

55. Pereira RMR, Siqueira N, Costa E, do Vale D, Alonso N. Unilateral cleft lip and palate surgical protocols and facial growth outcomes. J Craniofac Surg. 2018;29:1562–1568.

56. Xu X, Kwon HJ, Shi B, Zheng Q, Yin H, Li C. Influence of different palate repair protocols on facial growth in unilateral complete cleft lip and palate. J Craniomaxillofac Surg. 2015;43:43–47.

57. Fudalej P, Hortis-Dzierzbicka M, Dudkiewicz Z, Semb G. Dental arch relationship in children with complete unilateral cleft lip and palate following Warsaw (one-stage repair) and Oslo protocols. Cleft Palate Craniofac Jour. 2009;46:648–653.

58. Smyth AG, Knepil GJ. Prophylactic antibiotics and surgery for primary clefts. Br J Oral Maxillofac Surg. 2008;46:107–109.

59. Hupkens P, Lauret GJ, Dubelaar IJM, Hartman EHM, Spauwen PHM. Prevention of wound dehiscence in palatal surgery by preoperative identification of group A Streptococcus and Staphylococcus aureus. Eur J Plast Surg. 2007;29:321–325.

60. Aznar ML, Schönmeyr B, Echaniz G, Nebeker L, Wendby L, Campbell A. Role of Postoperative Antimicrobials in Cleft Palate Surgery: Prospective, Double-Blind, Randomized, Placebo-Controlled Clinical Study in India. Plast Reconstr Surg 2015;136(1):59e–66e.

61. Durga P, Raavula P, Gurajala I, et al. Evaluation of the efficacy of tranexamic acid on the surgical field in primary cleft palate surgery on children-A prospective, randomized clinical study. Cleft Palate Craniofac J. 2015;52:e183–e187.

62. Arantes GC, Pereira RMR, de Melo DB, Alonso N, Duarte M do CMB. Effectiveness of tranexamic acid for reducing intraoperative bleeding in palatoplasties: A randomized clinical trial. J Craniomaxillofac Surg. 2017;45:642–648.

63. Bateman MC, Conejero JA, Mooney EK, Rothkopf DM. Short-stay cleft palate surgery with intraoperative dexamethasone and Marcaine. Ann Plast Surg. 2006;57:245–247.

64. Senders CW, di Mauro SM, Brodie HA, Emery BE, Sykes JM. The efficacy of perioperative steroid therapy in pediatric primary palatoplasty. Cleft Palate Craniofac J. 1999;36:340–344.

65. Bouaggad A, Medraoui M, Bouderka MA. The effect of dexamethasone on recovery from cleft palate surgery. Eur J Anaesthesiol. 2007;24(1):99.

66. Henry C, Samson T, MacKay D. Evidence-based medicine: The cleft lip nasal deformity. Plast Reconstr Surg. 2014;133:1276–1288.

